# On the Spread of Coronavirus Infection. A Mechanistic Model to Rate Strategies for Disease Management

**DOI:** 10.1101/2020.06.05.20123356

**Authors:** Shiyan Wang, Doraiswami Ramkrishna

## Abstract

Effective policy making based on ongoing COVID-19 pandemic is an urgent issue. We present a mathematical model describing the viral infection dynamics, which reveals two transmissibility parameters influenced by the management strategies in the area for control of the current pandemic. The parameters readily yield the peak infection rate and means for flattening the curve. Model parameters are shown to be correlated to different management strategies by employing machine learning, enabling comparison of different strategies and suggesting timely alterations. Treatment of population data with the model shows that restricted non-essential business closure, school closing and strictures on mass gathering influence the spread of infection. While a rational strategy for initiation of an economic reboot would call for a wider perspective of the local economics, the model can speculate on its timing based on the status of the infection as reflected by its potential for an unacceptably renewed viral onslaught.

## Introduction

The pandemic of coronavirus (SARS-COV-2) infection has gripped the world with unparalleled anxiety. An alarming number of deaths have occurred within the short span of a little over four months! In US, more than one hundred thousand have died at the time of compiling this article with prospects of many more in the horizon. Despite the epidemic slowing, it appears to be abating at an unacceptable rate. There has been a scramble for controlling the spread of infection by people of various backgrounds including medical professionals, scientists, engineers, economists, the media, and political leaders. Although considerable insight has accumulated over efficient ways to confront this cataclysm *(1, 2)*, much more remains to be learned about the disease transmission, its treatment, and prevention by a suitable vaccine for the future. While the government has taken actions to relieve the economic burden of coronavirus on certain industries, businesses, and American workers (e.g., paycheck protection program), the looming prospects of an economic breakdown of catastrophic proportions are a further complication that must also somehow influence the mode of confrontation of the pandemic.

An essential prerequisite to facing the coronavirus pandemic is understanding of the various factors that have a potential contribution to limiting the spread. The spread of infection occurs in multifarious ways. Thus one that is cited the most frequently is spread of the virus through droplets from coughing and sneezing *(3)*. Another is from unwitting contact with infected surfaces *(4)* such as glassware, boxes and so on. Intimate contact through handshakes and hugs are even more efficient ways to transmit infection. Each occurs through different scenarios that must be envisaged with their respective frequencies of occurrence for a model formulation. For symptomatic disease associated with a pathogen transmissibility (marked by a basic reproduction number), different transmission routes are aligned to their implications for prevention; specifically, there may be four categories: symptomatic transmission, pre-symptomatic transmission, asymptomatic transmission, and environmental transmission. Given recent evidence of SARS-CoV-2 transmission by mildly symptomatic and asymptomatic persons *(5)*, its incubation period is about 5.1 days and about 12 days of infection from exposure to symptom development (latent period). Therefore, unusually long term of latency period and pre-symptomatic transmission could have important implications for transmission dynamics *(6)*.

Analysis of data accumulated from numerous sources have provided the general features of the spread in terms of when to expect the peak infection rate and what it takes to flatten this curve. Yet this understanding must be said to be qualitative without notable predictive features. A mathematical model is presented here of the spread of coronavirus (COVID-19) in terms of three parameters that control the rate of its spreading and flattening the infection rate curve when intervention by a vaccine is not available. Our model is concerned with a specific geographic domain of the United States with a given population of specified density (number per unit area) of which a fraction is initially infected. The infected population contributes virus within the domain which, for the present, is completely isolated from other domains. The spread of infection within the domain depends on the uninfected population and occurs at a rate governed by the extent of protective measures adopted to avoid infection from those infected. This spread obviously depends also on the viral population in the domain which grows by contribution from the infected (exhaled droplets, aerosol, contaminated surfaces, and possibly fecal-oral contamination *(7)*) and disappears by death/isolation/herd immunity etc. We should note that while there are numerous reports on reinfection of COVID-19 *(8)*, majority of recovered patients retain certain immunity against the virus.

Our goal here is to find a suitably simple framework to produce a mathematical model that contains a limited number of parameters which can be readily identified from gross observations. Furthermore, they should relate in some way to various strategies that may be envisaged to control the spread of infection. To simulate both dynamics of viral and infected population, the modeling of the system in a considered geometric domain can be abstracted as its dimensionless form

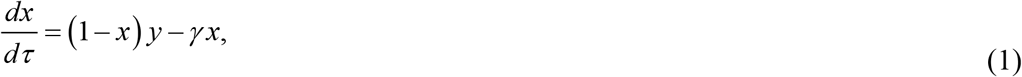

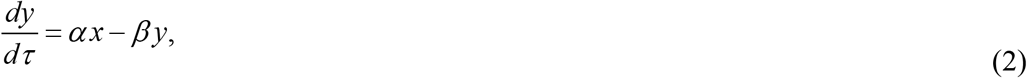

where *x* = *n* / *N*_*o*_ is the infected population density (*n*) normalized by the population density in the domain (*N*_*o*_), *y* = *V* / *V*_*o*_ is the dimensionless viral population density, *τ* = *t* / *T*_inf_ is the time scaled by the average time for an individual to be infected (*T*_inf_); The explanations of both dynamic equations are elaborated in supplementary material. Three dimensionless parameters (see physical interpretation of *α, β, γ* in Table 1) presented in above differential equations compare the rates of different processes and have the capacity to control the spread of infection. Daily infection data must be fitted to the model by appropriate choice for the values of the dimensionless parameters (see the model fitting in Fig. S1 in supplementary material). The socio-economic behavior has diversified the dynamics of the infection curve; Furthermore, major regulatory governmental strictures may enforce more discipline in public behavior thus seriously affecting the parameters. This effect, it must be conceded, is buried in subtle empiricism of the model that we must seek to unearth. In doing so, we emulate the currently popular practice of machine learning towards estimating the parameters in each domain to assess the local government policy. In this regard, the informative results delivered by combining both approaches (i.e. mechanistic model and machine learning) could promote effective policy implementations against the transmission of disease (Fig. 1(A)). In Fig. 1(B), the national scale social distancing is undertaken with the administration guideline “15 Days to Slow the Spread” that divides the pre-guideline enforcement period (P1) and the post-guideline enforcement period (P2). Furthermore, to consider the heterogeneity of the population density, we model the infection dynamics in the leading county of every state (50 states plus Washington D.C.) Within different periods and regions, their parameter values will reflect the quality of management of the spread of infection in the area under consideration. The different mechanisms of transmission of infection may operate to varying extents in different areas depending on how the infection is managed locally. Thus one must regard the model as only “broadly” mechanistic and the relationship of model parameters to different strategies would be somewhat diffuse. Therefore, in connecting the model to guide strategies we resort to a statistical methodology based on machine learning tools, which could overcome the limitation just mentioned.

**Table 1.** Emerging indicators from pathogen system.

| Notation | Variables |
| --- | --- |
| $\alpha \equiv \frac{k_v N_o}{k V_o^2}$ | Fractional rate of viral growth during the (average) infection time with $V_o$ viral population;<br>$k_v$ is the rate constant for production of virus from the infected;<br>$k$ is the rate constant for transfer of infection |
| $\beta \equiv \frac{k'_v}{k V_o}$ | Viral death rate during (average infection time);<br>$k'_v$ is the rate of death of virus |
| $\gamma \equiv \frac{k_r}{k V_o}$ | Removed rate of infected patients relative to infection rate with $V_o$ viral population;<br>$k_r$ is the rate of removed population (either by death or recovery) |
| $\frac{\alpha}{\beta} \equiv \frac{k_v N_o}{k'_v V_o}$ | Ratio of viral growth rate to its death rate |

**Fig. 1.**
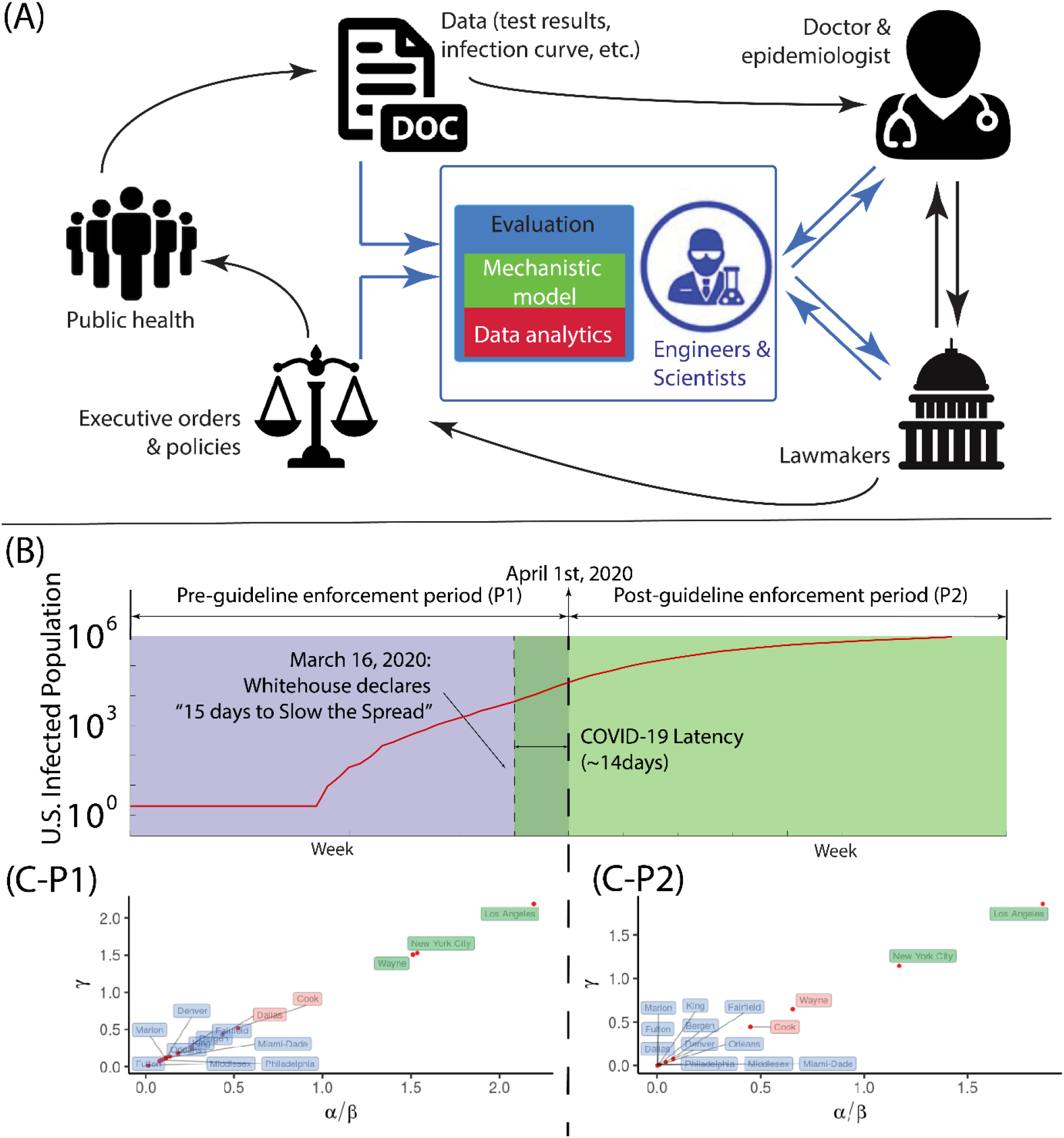
Parametric study on coronavirus infection in United States. (**A**) By incorporating both mechanistic modeling and data analysis (in blue) into the traditional workflow of the policy making (in black), a refreshed framework forms a three-way communication among expert (doctor/epidemiologist), engineers/scientists and lawmakers, thus improving the implementation of health policies against the infectious disease. (**B**) The timeline of total population with COVID-19 positive in conjunction with the policy of “15 Days to Slow the Spread” in the United States, in which the infection period is divided into the pre-guideline enforcement period (P1) and the post-guideline enforcement period (P2). (**C**) The phase space in terms of *α* / *β* and *γ* is plotted for the leading infected counties in first fifteen states: P1 duration and P2 duration. The phase space is segregated as three regions: ‘severe’ (labeled as green; *α* / *β* ∈(1.0, ∞)), ‘moderate’ (labeled as red; *α* / *β* ∈[0.4,1.0]), and ‘mild’ (labeled as blue; *α* / *β* ∈(0, 0.4)).

## Results

### Role of Parameters in Spread of Infection

We first examine role of model parameters in the spread of infection. The P1 duration reveals the period of pathogen transmission with limited prevention in the United States. The early state of virus transmissibility can be characterized by ‘R-naught’ (R0), which is the basic reproduction number. Our estimate of R0 is about 2.8 (the median from data is 2.75; our model is 2.90) whose transmission is stronger than influenza (R0:1.4-1.6) *(9)* and weaker than Measles (R0:12-18) *(10)*. The speed of infection of an individual would depend on the value of *T*_inf_ : a large *T*_inf_ would imply a longer real time and thus a slower rise in infection. For instance, in New York at P1 period without government policy intervention, it typically takes about *T*_inf_ ≈ 10 minutes to infect an individual. With the implementation of government policy about social distancing, in P2 period, *T*_inf_ increases 25 times, and the approximation of an individual infection takes about 4 hours! To measure parameters *(α, β, γ*), it is purposeful to examine the steady state solutions of the model represented by Eqs. (1) and (2). We show that the only relevant solution is 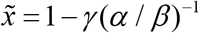 and 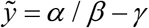 where the projected total infected population 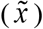 and viral population 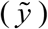 are determined by *α* / *β* and *γ*. Parameter *α* / *β* represents the ratio of viral growth rate to its death rate, which represents the extent of environmental virulence. Fig. 1(C) shows that severe virulence environment (large *α* / *β)* are associated with the large counties (i.e. Los Angeles-CA (P1,P2), New York City-NY(P1,P2)). In particular, Wayne county at Michigan State shows significant improvement (severe →moderate) as the proper social distancing is taken and thereby there would be a significant reduction of the virus in circulation. In general, counties with populous majority remain as small virulence during the entire period (Fig. 1(C)). Parameter *γ* represents the removal rate of infected patients (by recovery/death). Our model implies that *γ* is associated with *α* / *β* positively: despite the infection, its percentage in each county remains low (e.g. the percentage of infection at New York City is about *O*(10^−2^)); Therefore, *γ (α* / *β*)^−1^ ∼ 1 and always *γ* < *(α* / *β*). Here, we ought to demonstrate two significant points: (1) mathematically, *γ* < *(α* / *β*) is the necessary and sufficient condition for the stability of the solution; (2) as the difference between *γ* and *α* / *β* becomes smaller, the eventual infected population is smaller. The most direct way of containing infection depends on the availability of effective vaccines and therapies that can raise the value of *γ*. However, we should note that, without further modification, the current model would not directly account for the possibility of using experimental prescriptions such as Remdesivir recently authorized by Food and Drug Administration (FDA) *(11)*.

### Indicators of COVID-19 Transmissibility: *α* / *β* & 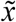

Based on our model, we propose two indicators *α* / *β* and 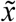 to characterize the infection dynamics. Reducing *α* / *β* is accomplished by slowing the viral transfer from the infected to the uninfected which can be accomplished by several ways such as social distancing (individual precautions can be washing, wearing masks, physical distancing 6 or 12 feet, and so on). In Fig. 2(A), we find that *α* / *β* is strongly correlated with the county population (R=0.91, p=2.1e-6; p<0.05 considered as significant), which is consistent with the physical explanation of *(α* / *β*) ∝ *(N*_*o*_ / *V*_*o*_) in Table 1; given any domain, *(N*_*o*_ / *V*_*o*_) increases with large population number but independent of population density *N*_*o*_. On the other hand, the projected total infection fraction 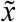, which characterizes the pathogen transmissibility, is positively correlated with the county population density (R=0.61, p=0.016 in Fig. 2(B)) because of *N*_*o*_ ; the transmission of the infection becomes faster when the population density is high. Other factors such as weather (temperature, humidity) remain insignificant (weak correlation) to the infection dynamics (see supplementary material; also (12)). Fig. 2(C) exhibits how parameters *α* / *β* and 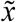 affect the peak infection rate. The peak infection rate represents the stage beyond which the infection rate will drop. It is now possible to address the much debated strategy of “flattening the curve” by lowering peak infection rate. To reduce the transmissibility (i.e. lower 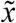), the peak infection rate has to be small (see Fig. 2(C)). Our model recommends that this is accomplished when *α* / *β* is low, suggesting the reduction of virus circulation (Fig. 2(D)).

**Fig. 2.**
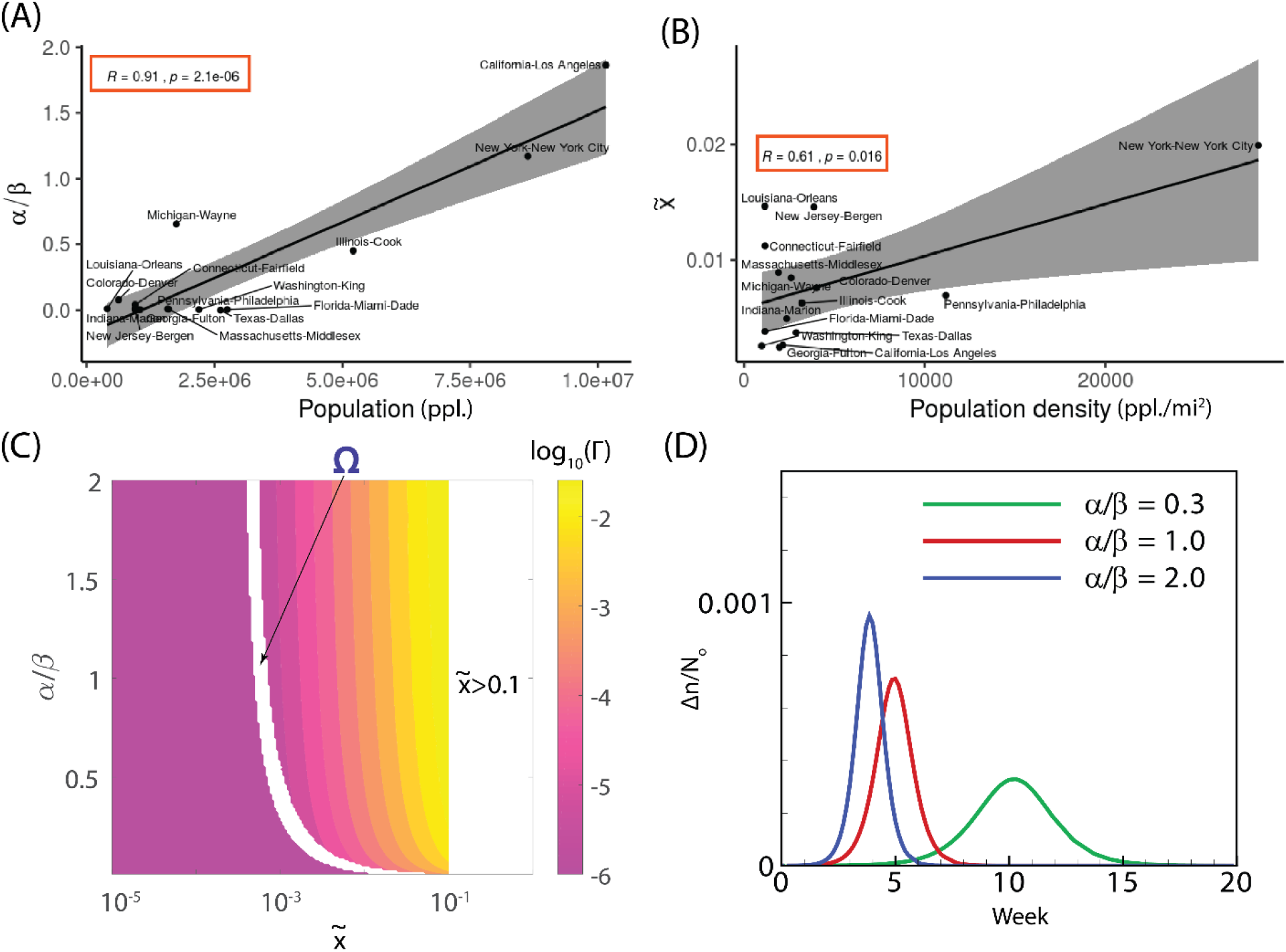
Relationship between county population and *α* / *β* & 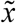. (**A** and **B**) The statistical correlations *(R*: Pearson correlation coefficient) are displayed for (A) virulence *(α* / *β)* vs. the corresponding county population and (B) the projected infection fraction 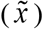 vs. the corresponding county population density (sample number is 15). (**C**) A phase diagram in terms of *α* / *β* and 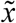 for the peak infection rate Γ ; domain ‘Ω’ represents the region in which the peak infection is not reached and Region 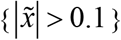 indicates the unlikely space where total infection exceeds 10% of the total population in the domain considered. (**D**) The effect of dimensionless parameter *α* / *β* on peak infection is studied with 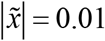. For (C) and (D), the simulations are conducted for a duration of 1 year (365 days) with *T*_inf_ = 40 minutes.

### Impact of Lockdown: New York City

We now proceed to model the effect of lockdown on COVID-19 transmissibility in New York City, as an example. Additionally, considering recently published policy of “Opening Up America Again” by the white house administration, we will study the effect of reopening economy on the dynamics of transmissibility in the New York metropolitan area. Here, we consider the influence of lockdown policy at New York City, where the isolation is determined by the geographic constraint of five boroughs (the Bronx, Brooklyn, Manhattan, Queens, and Staten Island) within the New York City (Fig. 3(A)). From Fig. 3(B), our model suggests that the mitigation brought about by lockdown is sensitive to the moment of implementation; an early enforcement of lockdown could delay its occurrence. Our study also shows that if the implementation happens after the peak infection, the strategy of slowing works less efficiently. However, the lockdown likely results in a long-term dynamics and the associated economic damage has to be considered as well.

**Fig. 3.**
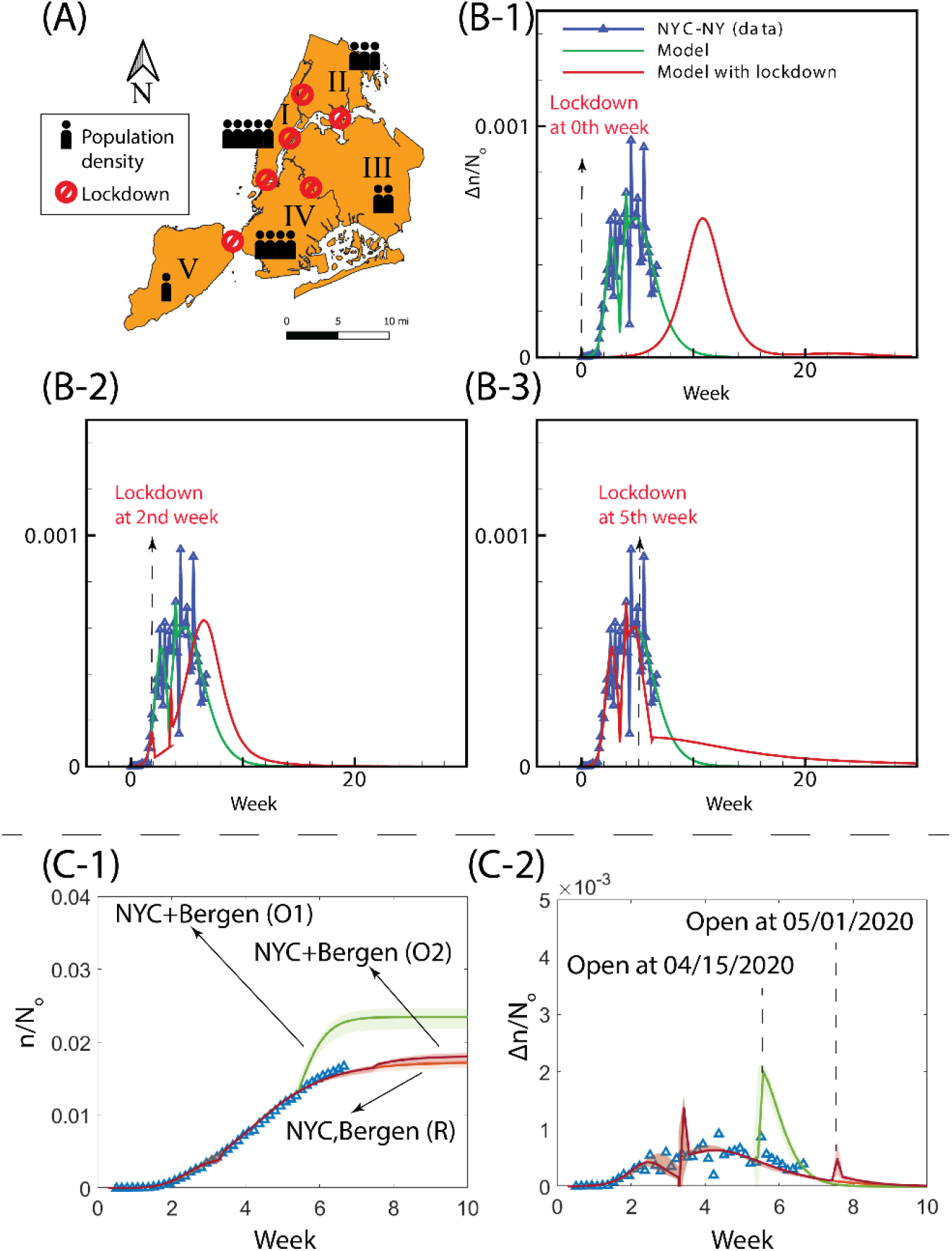
Effect of lockdown and reopening economy on the spread of infection in New York City. (**A**) The strict policy of lockdown is assumed at the borders of five boroughs - Manhattan(I), the Bronx (II), Queens (III), Brooklyn (IV) and Staten Island (V) within New York City. (**B**) The date of initiating lockdown affects daily positive cases Δ*n* in New York City scaled by total population density *N*_*o*_, where the start of policy enforcement is set at different initiation points: zeroth week (B-1), 2nd week (B-2), and 5th week (B-2), respectively. (**C**) Modeling of infected population (C-1) and increment (C-2) by considering different opening periods for New York City and Bergen county (inner New Jersey circle): ‘NYC+Bergen (O1)’ indicates the reopening economy at 5.5th week; ‘NYC+Bergen (O2)’ indicates the reopening economy at 7.5th week; ‘NYC, Bergen (R)’ indicates the economy remains closed. In (C), *N*_*o*_ represents the averaged resident population density of both New York City and Bergen (New Jersey); Symbols ‘Δ’ are the data of the infected population within New York City and Bergen (New Jersey); Plots with shaded area are the modeling results where solid lines represent the median of the prediction and the shaded area indicates the uncertainty. The zeroth week is set at the moment when the total number of infections at the New York City is ten.

### Reopening Economy on Second Wave: New York Metro Area

During this pandemic, the financial center of the world--New York City area has been hit by massive layoffs and anticipates looming recession (13). This situation, spells some urgency for reopening the economy and resuming normal daily activity. However, we stress that opening the economy has to be cautious to the possible appearance of a second wave thus making the timing of the reopening very important. To simulate the impact of normal daily activity on the current dynamics of infection, we study the transmissibility in both New York City and Bergen County (Inner New Jersey) within the metropolitan area. These two regions represent the most active interactions in the United States (leading out-computing in the metro area, NYC Planning 2018) and yet both have the leading coronavirus infections in their states. In the model, we relax the current government restraints and resume normal daily operations and activities, which allows the model to consider the worst scenario of the infection curve. Fig. 3(C) show that the economy reopening (with the least precaution) inevitably brings the second wave and thereof more mortality. However, the extent of infection outbreak can be drastically reduced by delaying the opening date (35% increase at 5.5th week vs. 4% increase at 7.5th week). We note that an effective policy intervention may reduce the drastic increment of the infected population. In the next section, we discuss how to quantify the effectiveness of current implemented policy on coronavirus transmissibility.

### Influence of Policy Measures in States

With the U.S. administration declaring the social distancing guideline since March 16th, local governments have implemented more than 300 executive orders in fifty states, Puerto Rico, the District of Columbia, Guam, and the Virgin Islands. The executive actions and policies are related to declarations of states of emergency, school/business closure, prohibition of mass gathering, stay at home order, etc. Central issues stand as the effectiveness of ongoing individual policy is unclear. In our study, while the coronavirus transmissibility is strongly influenced by the local population dynamics (Figs. 2(A, B)), the statistical inference suggests the relevance of several ongoing policies on the coronavirus transmission (Fig. 4(A)). With our model empowered by machine learning tools, we perform the regression of both parameters *α* / *β* and 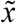 based on all policy influences. By examining the weight associated with each policy measure and its significance (p-value) in Fig. 4(B-1), we should conclude that factors such as non-essential business closure, gathering ban and school closure possess strong impact on 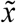 (adjusted-R2=0.59, p=2e-6), which represents the total infected population. Both gathering ban and school closure emphasize the activity of population in young age, which is consistent with the recent finding that young people play a vital role in spread of COVID-19 (14,15). For virulence environment *(α* / *β*), while the severity of coronavirus spread is largely determined by the local population number, nonessential business closure plays a role in its attenuation effort among other considered policies (adjusted-R2=0.30, p=1e-3; see Fig. 4(B-2)). With the context of reopening the economy (since May 1st 2020), there are several reports on the surge of coronavirus infection in multiple states and most contagious region remains at the leading counties. Policies on both certain non-essential business limitation and gathering ban have been revised in almost all states (note: schools remain closed across the United States). Our model enables to incorporate these policy changes to predict and diagnose many states having surges of positive cases (Fig. 4(C)). By predicting the increase of *α* / *β* and 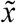 using machine learning, we find that the strong surges (marked as ‘severe’ in Fig. 4(C)) in states like Utah, Nebraska, Ohio, Kentucky, Texas and Virginia could stem from the relaxation of the gathering ban; Nevada, North Carolina, South Carolina and Mississippi have observed high daily positive cases, which is interpreted by our model as due to the non-essential businesses; We also find that several states (e.g. California, Wisconsin, Arizona, Alaska, Tennessee, Maine, etc.) have strong infection due to both factors (non-essential business closure and gathering ban). Our model captures all currently emerging states, indicating significant impact of government policy on the spread of coronavirus when a vaccine is unavailable. In Fig. 4(C), several states (marked as ‘moderate’) should be cautiously optimistic when relaxing social distancing measures despite downward trending of daily positive number because we observe increases in either *α* / *β* (elevated virulence) or 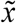 (increased projected total infection number) in our model. In particular, the state of Massachusetts should retain strong measures (increments in both *α* / *β* and 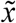).

**Fig. 4.**
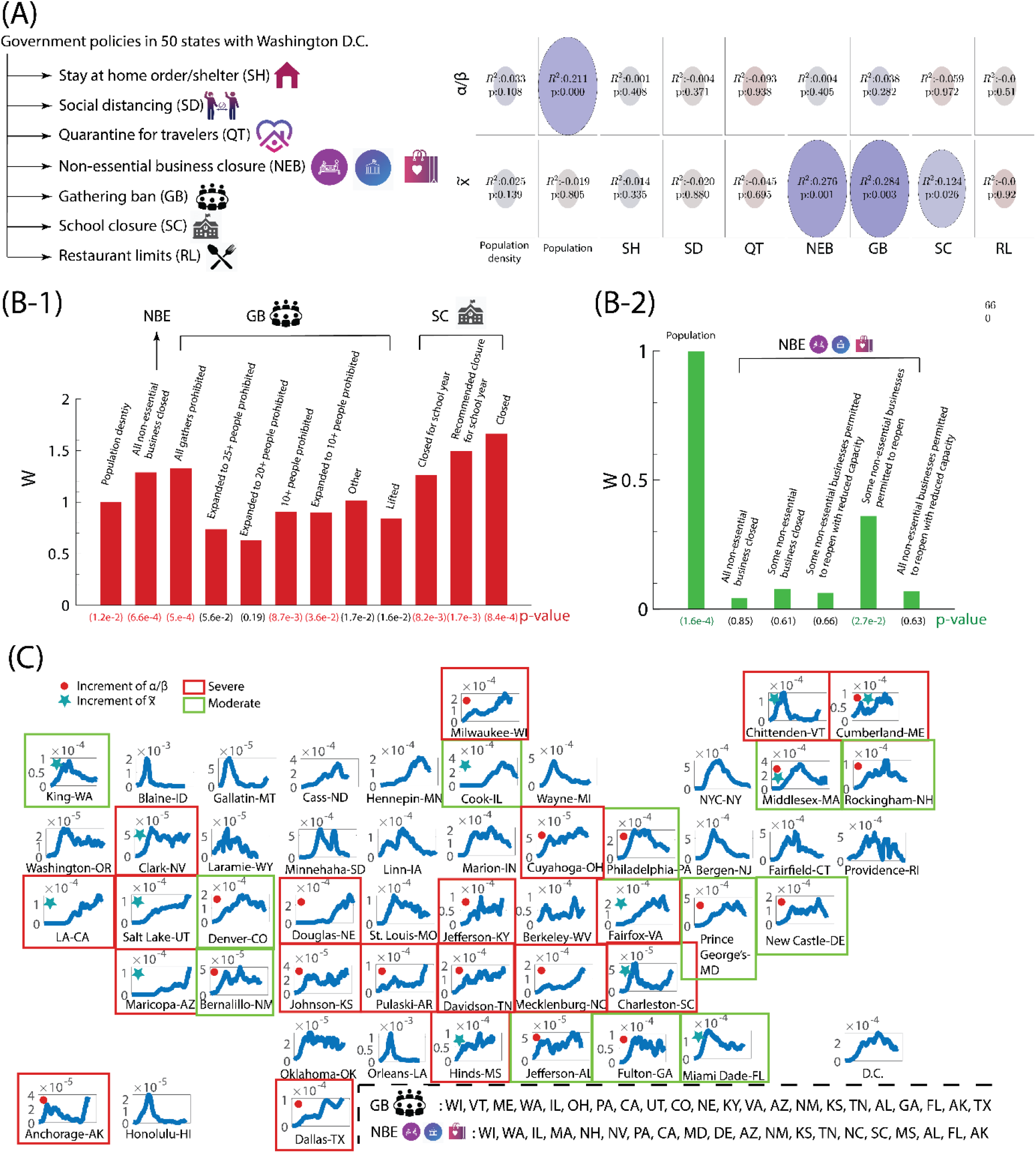
Evaluation of policies on COVID-19 transmissibility. (**A**) The relevance and significance of individual policy and population dynamics (population and population density) on model parameters *α* / *β* and 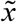, characterized by adjusted R-square and p-values, respectively. In the diagram, the color of the ellipse represents the value of adjusted *R*^*2*^ while the size of ellipse accounts for p-value. (**B**) The regression analysis of government policies and local population dynamics against (B-1) the projected eventual infection fraction 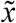 and (B-2) virulence *α* / *β*. The extent of influence from individual factor is reflected by the corresponding weight *(W)*. In (B), the weights are normalized by the first factor. (**C**) Upon economy opening, the modification of polices impacts the trend of infection curve. We can identify the states with the surge of positive cases by the prediction of *α* / *β* and 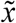 using machine learning: the blue plots represent the daily increment Δ*n* (7-day running averaged) normalized by the total population density *N*_*o*_ in the leading county of all fifty states (including D.C.) For all ‘emerging’ counties in each state, a red filled circle is recognized by the elevation of *α* / *β* from the prediction of the machine learning; A green filled star represents the increase of 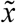; the red square implies the severe situation based on the plateau/ upward-trending daily growth and the yellow square indicates the moderate scenario based on the downward-trending daily growth curve.

## Discussion

In this report, we have proposed a new mechanistic model describing the transmission of COVID-19 in the United States. Our model is established in conjunction with administration policy, from which we propose two significant parameters. The parameter *α* / *β* quantifies the severity of the coronavirus circulation, and the parameter 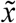 represents the projected total infected fraction. To be consistent with CDC county-by-county guideline, we studied the infection dynamics of the leading county in each state. Our study shows that New York City in New York, Los Angeles county in California, and Wayne county in Michigan exhibit strong coronavirus circulation. By examining the peak infection rate, our suggested strategy of ‘flattening the curve’ has to deal with lowering *α* / *β*, towards drastically diminishing the virus population in the environment. Our study of lockdown suggests that it be implemented *before* the peak infection rate, since its arrival can be sensed well by the parameters. We have quantified the impact of current social distancing policies with *α* / *β* and 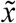, suggesting that polices such as, restrictive non-essential business closure, a ban on gathering, and that of school closure are critical. This may strongly associate with the restricted activity of young people (young adults and teenagers). The novelty of this contribution is derived from two specific features. One locates each geographic region in our parameter space at any stage providing for a diagnosis of the disease status in the region, and the prevailing quality of its management. The other affords a direction in which changes in strategy must be brought about for controlling the disease. Although a rational analysis for an economic reboot should be based on a considerably expanded view of the local economics, it is possible to derive some useful guidelines from our model study. To this extent, we conclude, perhaps somewhat speculatively, that our suggestion for an economic reopening may be viable if non-essential business closure is conditional, mass gathering is limited and school opening is delayed. At any rate, in the absence of such restrictive measures, the prospect of an economic recovery is less likely.

## Data Availability

The data that support the findings of this study are available from the corresponding author upon reasonable request.

## Author contributions

Conceptualization: SW, DR; Data curation: SW, DR; Formal analysis: SW, DR; Methodology: SW, DR; Software: SW, DR; Visualization: SW, DR; Writing, original draft: SW, DR; Writing, reviewing and editing: SW, DR.

## Data and materials availability

US coronavirus data are publically available from the New York Times GitHub source: https://raw.githubusercontent.com/nytimes/covid-19-data/master/us-counties.csv; Weather data are publicly available from NOAA Global Surface Summary of the Day (GSOD): https://data.nodc.noaa.gov/cgi-bin/iso?id=gov.noaa.ncdc:C00516; State and policy data to address coronavirus are publically available from Kaiser Family Foundation: https://www.kff.org/health-costs/issue-brief/state-data-and-policy-actions-to-address-coronavirus/

